# Associations between vitamin D and autoimmune diseases: Mendelian randomization analysis

**DOI:** 10.1101/2023.04.08.23288323

**Authors:** Sizheng Steven Zhao, Amy Mason, Eva Gjekmarkaj, Haruyuki Yanaoka, Stephen Burgess

## Abstract

**Objective:** The VITAL trial of vitamin D supplementation suggested a possible protective effect for autoimmune diseases but uncertainties remain. We investigated potential causal effects of vitamin D on composite and individual autoimmune diseases using Mendelian randomization.

**Methods:** We used data from 332,984 participants of the UK Biobank of whom 23,089 had at least one autoimmune disease defined using ICD code and/or self-report. Diseases were further considered in mechanistic subgroups driven by “autoimmunity” (n=12,774) or “autoinflammation” (n=11,164), then individually. We selected variants within gene regions implicated in vitamin D biology to generate a weighted genetic score. We performed population-wide analysis using the ratio method, then examined non-linear effects across five quantiles based on 25-hydroxycholecalciferol levels.

**Results:** Genetically-predicted vitamin D was associated with lower risk of diseases in the autoinflammation group (OR 0.95 per 10ng/ml increase in 25-hydroxycholecalciferol; 95%CI 0.91-0.99; p=0.03) but not the autoimmunity group (OR 0.99; 95%CI 0.95-1.03; p=0.64) or combined. When considering individual diseases, genetically-predicted vitamin D was associated with lower risk of psoriasis (OR 0.91; 95%CI 0.85-0.97; p=0.005), the most common disease in the autoinflammation group, and suggestively with systemic lupus erythematosus (OR 0.84; 95%CI 0.69-1.02; p=0.08); results were replicated using data from independent studies. We found no evidence for a plausible non-linear relationship between vitamin D and any outcome.

**Conclusions:** We found genetic evidence to support a causal link between 25-hydroxycholecalciferol concentrations and psoriasis and systemic lupus erythematosus. These results have implications for potential disease prevention strategies, and the interpretation and design of vitamin D supplementation trials.

## INTRODUCTION

Autoimmune diseases are a heterogenous group of conditions that are among the leading causes of life-changing morbidity and even mortality [1,2]. Pharmacological therapies that target the immune system are not always effective and can have prohibitive adverse effects. Vitamin D is highly popular as a complementary and alternative medicine. Enthusiasm for it as a potentially disease modifying agent has largely been driven by pre-clinical studies and a host of observational associations that are at risk of bias from confounding (e.g., factors such as lifestyle and diet that influence both vitamin D levels and disease risk) and reverse causation (e.g., reduced sun exposure and/or dietary absorption due to the autoimmune disease) [3].

Adequately powered randomised control trials of vitamin D supplementation among people with autoimmune diseases are scarce. In the recent VITAL trial [4], vitamin D supplementation (2000 IU/day over a median of 5.3 years) reduced risk of autoimmune diseases as a composite outcome (hazard ratio 0.78, 95% confidence interval 0.61-0.99, P=0.05). However, uncertainties remain in part due to a relatively small number of events (n=278 among 25,871 participants), possibly because the mean age of the trial population (67 years) is older than the peak incidence age for some autoimmune diseases. The trial was not powered to examine individual autoimmune diseases, which is important since any biologic effects of vitamin D on the immune system is unlikely to be shared across pathologically diverse conditions. VITAL was also not powered to examine whether risk reduction differed according to baseline vitamin D, which is important to assess potential threshold or non-linear effects of intervention.

Genetic instrumental variable designs, also known as Mendelian randomization (MR), can help with these challenges. Since genetic variants are randomly allocated at conception, MR is typically more robust against confounding and reverse causation compared to traditional observational designs. Large population level genetic data can power subgroup analyses of pathologically related conditions, while recent methodologic developments can help examine non-linear effects of vitamin D interventions. We conducted both population-wide and stratified MR analyses to assess evidence for potential causal effects of vitamin D on autoimmune diseases and to interrogate any non-linearity in the relationship.

## METHODS

### Study populations and outcomes

We performed population-wide and stratified Mendelian randomization analyses in the UK Biobank, a prospective cohort study of around 0.5 million participants aged 40 to 69 years at baseline, recruited between 2006-2010 in the United Kingdom and followed-up for a median of 10.9 years [5]. UK Biobank has approval from the North West Multi-centre Research Ethics Committee and all participants provided written informed consent. The UK Biobank received ethical approval Analyses were restricted to unrelated individuals of European ancestries who passed various quality control steps as previously described [6] and had a valid 25-hydroxycholecalciferol (25(OH)D) measurement.

We considered a predefined list of outcomes based on ICD-9 and -10 codes (from fields 41270, 40001 or 40002) and/or self-reported diagnosis (field 20002); the full list of conditions, sample size and definitions are shown in **Supplementary Table 1**. The conditions were first studied as a composite of “all autoimmune diseases”, excluding multiple sclerosis (MS) for which prior MR studies have suggested a causal relationship with vitamin D [7]. We then broadly classified diseases in two groups according to a proposed classification method based on shared pathology and clinical phenotype [8]:

1. Disease driven by “autoimmunity” (i.e., aberrant dendritic and adaptive immune cell responses leading to breaking of tolerance and immune reactivity towards native antigens [8]): rheumatoid arthritis (RA), systemic lupus erythematosus (SLE), Sjögren’s syndrome, systemic sclerosis, Graves’ disease, Hashimoto’s thyroiditis, Coeliac disease, type 1 diabetes mellitus, primary biliary cholangitis, autoimmune hepatitis, polymyalgia rheumatica (PMR), giant cell arteritis (GCA), polyarteritis nodosa, Henoch-Schönlein purpura, granulomatosis with polyangiitis, eosinophilic granulomatosis with polyangiitis, microscopic polyangiitis, mixed connective tissue disease, antiphospholipid syndrome, dermatomyositis and polymyositis.
2. Disease driven by “autoinflammation” (i.e., local factors at sites predisposed to disease lead to innate immune cell activation with resultant target tissue damage [8]): ankylosing spondylitis, psoriatic arthritis, psoriasis, Crohn’s disease, ulcerative colitis, primary sclerosing cholangitis, Behcet’s disease, Takayasu arteritis and Kawasaki disease.

Where mechanisms were less clear, diseases that were male-predominant and/or associated with HLA-B genes were grouped into the “autoinflammation” group [9] and remainder to the “autoimmunity” group. In sensitivity analyses, we limited analyses to diseases with >100 cases and for which the mechanism is better characterised within this classification system (i.e., the first ten in the autoimmunity group and first five in the autoinflammation group). It is possible for individuals to have more than one disease that belong to both groups. In further sensitivity analyses, we also investigated outcomes defined as those only with diseases in the autoimmunity or autoinflammation group.

We then analysed individual disease that affected at least 100 participants, since it was unlikely that smaller sample sizes would provide meaningfully powered analyses. We included participants with multiple autoimmune diseases for analyses of individual diseases, since some diseases commonly co-exist (e.g., ankylosing spondylitis and psoriasis).

For all analyses, controls comprised those without any of the above conditions or MS. We recognise that the control group will include individuals with other, rarer autoimmune conditions, but both absolute numbers and proportions are small.

We included MS as a positive control outcome [7], and osteoarthritis, which is not considered an autoimmune disease, as a negative control outcome.

### Vitamin D measurement and classification

Concentrations of 25(OH)D in blood were measured using the DiaSorin Liaison immunoassay analyser. Measurements were adjusted for month of blood draw to correspond to a measurement taken in October by subtracting the mean 25(OH)D concentration for the month the measurement was taken in and then adding the mean 25(OH)D concentration measurements taken in October. October was chosen as the reference month as 25(OH)D concentrations were close to their average annual value in October (**Supplementary Table 2**).

### Genetic variants

To minimize potential bias due to horizontal pleiotropy, we considered genetic variants from four gene regions previously shown to be strongly associated with 25(OH)D [10] and implicated in the transport, metabolism, and synthesis of vitamin D – *GC, DHCR7, CYP2R1*, and *CYP24A1*. To maximize the variance explained by the genetic instrument, we created a weighted genetic score from 21 variants associated with 25(OH)D concentrations at each genetic locus selected using a stepwise selection method (**Supplementary Table 3**). A prior study showed that this genetic risk score was not associated with potential risk factors for autoimmune diseases in UK Biobank, except for BMI, although this association was small [10]. By contrast, a score using variants from across the genome-wide score was strongly associated with other traits that may introduce bias from pleiotropy.

### Statistical methods

Population-wide analyses were performed by calculating the ratio between the association of the genetic score with the outcome and the association of the score with 25(OH)D concentrations. We performed logistic regression to estimate the associations of the score with the outcomes adjusting for age, sex, assessment centre and 10 genetic principal components of ancestry. MR estimates were scaled to a 10 nmol/L increase in genetically predicted 25(OH)D level.

In non-linear stratified analyses, we divided participants into five quantiles using the doubly ranked method [11]. We firstly divide the population into pre-strata based on the instrument level, and then divide into final strata based on the exposure level within each pre-stratum. Use of the doubly-ranked method is important for 25(OH)D as an exposure, as genetic associations with 25(OH)D levels vary strongly in the population [12]. MR estimates were calculated within each stratum as in the population-wide analyses, but using genetic associations estimated in each stratum of the population in turn.

### Supplementary analyses

Sample sizes for individual autoimmune diseases are typically smaller in the UK Biobank compared to dedicated GWAS consortia, which may limit the power of one-sample MR. Where possible, we sought to replicate suggestive causal associations (arbitrarily defined as p value<0.1) between 25(OH)D and individual diseases using two-sample MR. Outcome genetic data were taken from GWAS of psoriasis (10,588 physician diagnosed cases and 22,806 controls) and SLE (5,201 physician diagnosed cases and 9,066 controls). Where such GWAS data were not available, we attempted to replicate using data from FinnGen Release 8 [13] for GCA (884 cases, 332,115 controls) and PMR (3039 cases, 332,115 controls). Instruments were identified as genome-wide significant (p<5×10-8) and uncorrelated (r2<0.01) variants within *GC, DHCR7, CYP2R1*, and *CYP24A1* genes as above, taken from a GWAS meta-analysis of 25(OH)D [14]. We used the inverse variance weighted method which combines ratio estimates from each variant and, where possible, pleiotropy robust sensitivity analyses (i.e., MR Egger and weighted median/mode methods).

## RESULTS

Of 332,984 participants included for analysis, 23,089 had one or more autoimmune diseases. Their mean age was 59 years and 59% were females. The mean 25(OH)D concentration was 49 nmol/L, and similar in both autoimmune disease and control groups (**Table 1**). 12,774 had at least one disease of the “autoimmunity” group, and 11,164 had at least one disease in the “autoinflammation” group. There was a higher proportion of females in the autoimmunity than the autoinflammation group (69 vs 47%).

**Table 1.** Characteristics of autoimmune disease group and subgroups.

|  | All autoimmune diseases | Autoimmunity subgroup | Autoinflammation subgroup | Controls |
| --- | --- | --- | --- | --- |
| N | 23089 | 12774 | 11164 | 308521 |
| Age, years | 58.7 (7.8) | 59.7 (7.4) | 57.6 (7.9) | 57.0 (8.1) |
| Females | 13584 (58.8) | 8853 (69.3) | 5269 (47.2) | 163153 (52.9) |
| BMI | 27.8 (5.1) | 27.7 (5.3) | 27.9 (5.0) | 27.3 (4.7) |
| Vitamin D, nmol/L | 49.3 (21.7) | 50.0 (22.0) | 48.5 (21.4) | 49.4 (20.8) |
| Rheumatoid arthritis | 5172 (46.3) | 5172 (40.5) | 436 (3.9) | 0 |
| Systemic lupus erythematosus | 527 (4.7) | 527 (4.1) | 36 (0.3) | 0 |
| Systemic sclerosis | 201 (1.8) | 201 (1.6) | 11 (0.1) | 0 |
| Sjögren's syndrome | 335 (3) | 335 (2.6) | 23 (0.2) | 0 |
| Graves' disease | 1798 (16.1) | 1798 (14.1) | 85 (0.8) | 0 |
| Hashimoto's thyroiditis | 194 (1.7) | 194 (1.5) | 6 (0.1) | 0 |
| Coeliac disease | 2249 (20.1) | 2249 (17.6) | 125 (1.1) | 0 |
| Type 1 diabetes mellitus | 341 (3.1) | 341 (2.7) | 12 (0.1) | 0 |
| Primary biliary cholangitis | 243 (2.2) | 243 (1.9) | 35 (0.3) | 0 |
| Autoimmune hepatitis | 153 (1.4) | 153 (1.2) | 19 (0.2) | 0 |
| Polymyalgia rheumatica | 1780 (15.9) | 1780 (13.9) | 90 (0.8) | 0 |
| Giant cell arteritis | 436 (3.9) | 436 (3.4) | 20 (0.2) | 0 |
| polyarteritis nodosa | 91 (0.8) | 91 (0.7) | 10 (0.1) | 0 |
| Granulomatosis with polyangiitis | 62 (0.6) | 62 (0.5) | 7 (0.1) | 0 |
| Henoch-Schonlein purpura | 47 (0.4) | 47 (0.4) | 7 (0.1) | 0 |
| Eosinophilic Granulomatosis with polyangiitis | 46 (0.4) | 46 (0.4) | 7 (0.1) | 0 |
| Mixed connective tissue disease | 41 (0.4) | 41 (0.3) | 4 (0.04) | 0 |
| Antiphospholipid syndrome | 28 (0.3) | 28 (0.2) | 2 (0.02) | 0 |
| Microscopic polyangiitis | 19 (0.2) | 19 (0.1) | 1 (0.01) | 0 |
| Dermato/polymyositis | 6 (0.1) | 6 (0.05) | 0 | 0 |
| Ankylosing spondylitis | 1121 (10.0) | 111 (0.9) | 1121 (10.0) | 0 |
| Psoriatic arthritis | 703 (6.3) | 89 (0.7) | 703 (6.3) | 0 |
| Crohn's disease | 1795 (16.1) | 162 (1.3) | 1795 (16.1) | 0 |
| Ulcerative colitis | 3461 (31.0) | 278 (2.2) | 3461 (31.0) | 0 |
| Psoriasis | 5120 (45.9) | 335 (2.6) | 5120 (45.9) | 0 |
| Primary sclerosing cholangitis | 35 (0.3) | 16 (0.1) | 35 (0.3) | 0 |
| Behçet's disease | 30 (0.3) | 7 (0.1) | 30 (0.3) | 0 |
| Takayasu arteritis | 6 (0.1) | 1 (0.01) | 6 (0.1) | 0 |
| Kawasaki disease | 1 (0.01) | 1 (0.01) | 1 (0.01) | 0 |

2 Values represent mean (standard deviation) for continuous traits, and number of cases (percentage 3 of total cases) for binary traits and disease outcomes. Autoimmune diseases were separated into 4 those characterised by autoimmunity (i.e., aberrant dendritic and adaptive immune cell responses 5 leading to breaking of tolerance and immune reactivity towards native antigens) and 6 autoinflammation subgroups (i.e., local factors at sites predisposed to disease lead to innate 7 immune cell activation with resultant target tissue damage).

The genetic risk score explained 4.7% of the variance in 25(OH)D concentrations. There was some evidence that genetically predicted 25(OH)D was associated with reduced risk of autoimmune diseases overall, but confidence intervals included the null (OR 0.97 per 10 nmol/L increase in 25(OH)D; 95%CI 0.94, 1.01, p=0.10). This association appears to be predominantly driven by diseases in the autoinflammation group (OR 0.95; 95%CI 0.91, 0.99; p=0.03) (**Figure 1**). Sensitivity analyses showed similar results when restricted to ten autoimmunity and five autoinflammation diseases with >100 cases and for which disease mechanism is better characterised. Results were also similar after excluding individuals with diseases in both subgroups showed similar estimates.

**Figure 1.**
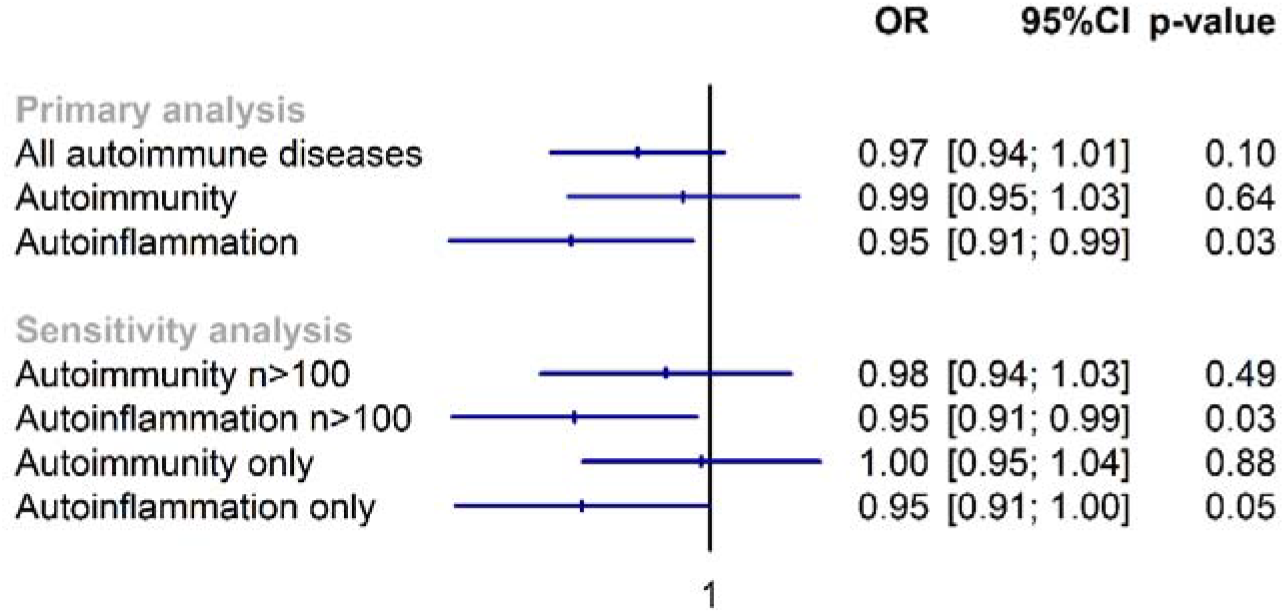
Association between genetically predicted vitamin D concentration and autoimmune diseases, including autoimmunity and autoinflammatory subgroups. Individuals in the primary analysis could have diseases in both autoimmunity (i.e., aberrant dendritic and adaptive immune cell responses leading to breaking of tolerance and immune reactivity towards native antigens) and autoinflammation subgroups (i.e., local factors at sites predisposed to disease lead to innate immune cell activation with resultant target tissue damage). The first set of sensitivity analyses were restricted to diseases with >100 cases for which disease mechanisms are better understood. In the second set of sensitive analyses, individuals with disease belong to both groups were excluded.

When diseases (with n>100) were analysed individually, psoriasis (OR 0.91; 95%CI 0.85, 0.97; p=0.005), GCA (OR 0.79; 95%CI 0.64, 0.98; p=0.03), PMR (OR 1.12; 95%CI 0.997, 1.25; p=0.06) and SLE (OR 0.84; 95%CI 0.69, 1.02; p=0.09) showed some evidence of association with 25(OH)D concentration (**Figure 2**). Genetically predicted 25(OH)D was not associated with the negative control, osteoarthritis, but was associated with lower risk of the positive control, MS.

**Figure 2.**
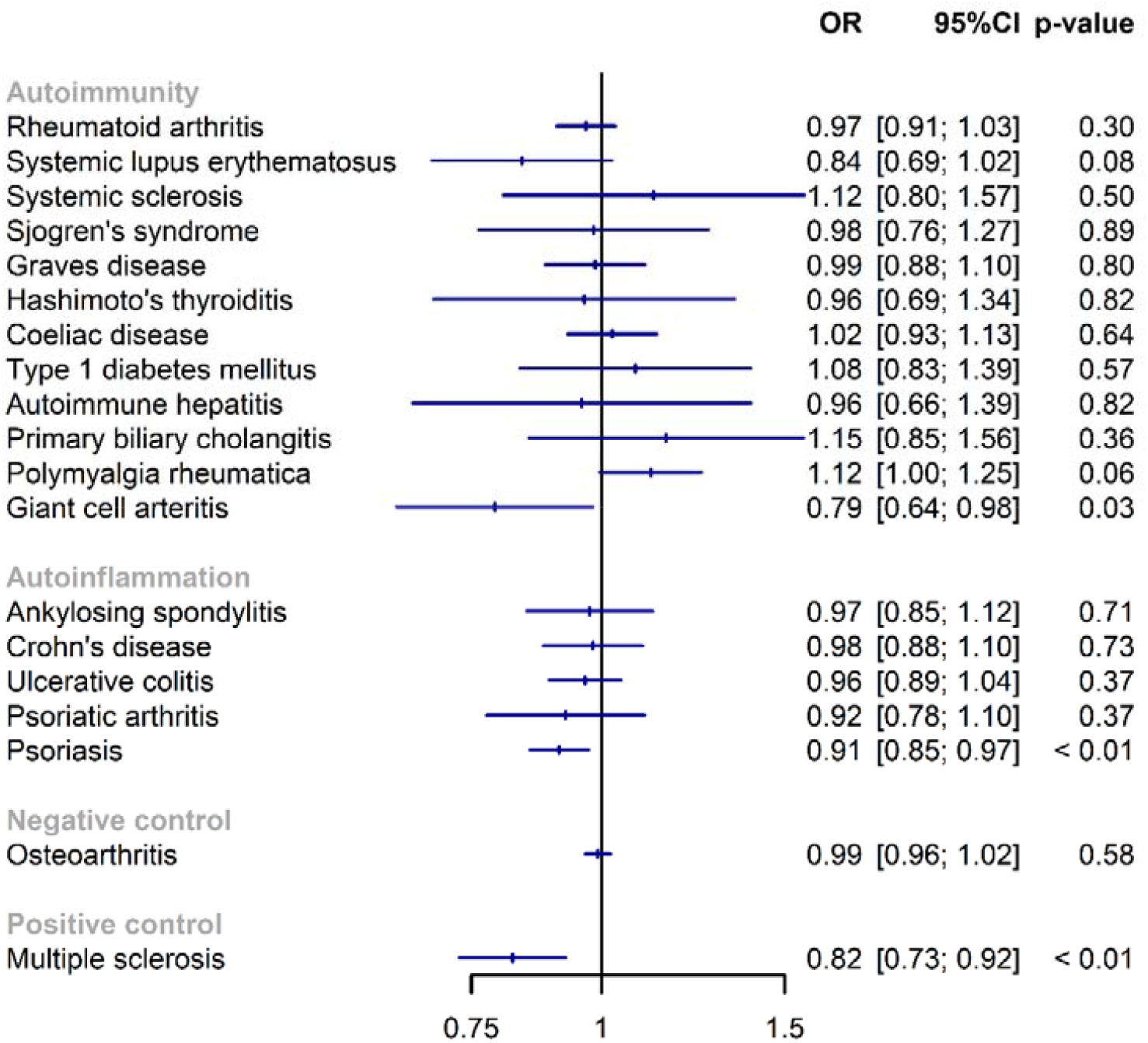
Associations between genetically predicted vitamin D concentration and individual autoimmune diseases.

There was statistical evidence for non-linear associations, albeit with limited biologic plausibility, between genetically-predicted 25(OH)D and the autoinflammation subgroup in the second (OR 0.82; 95%CI 0.68, 0.995; p=0.04) and fourth quantiles (OR 0.77; 95%CI 0.63, 0.93; p=0.006), but not any other quantiles (**Supplementary Table 4**).

For psoriasis, GCA, PMR and SLE, we attempted to replicate analyses using two-sample MR. 14 variants were used to instrument 25(OH)D. Genetically-predicted 25(OH)D was associated with lower risk of psoriasis (OR 0.52 per unit increase in log-transformed 25(OH)D; 95% 0.28, 0.96; p=0.04) and SLE (OR 0.61; 95%CI 0.40, 0.93; p=0.02). Estimates were directionally concordant in pleiotropy robust sensitivity analyses (**Supplementary Table 5**). Primary analysis results were not replicated for GCA (OR 0.95; 95%CI 0.47, 1.90; p=0.88) or PMR (OR 0.97; 95%CI 0.71, 1.33; p=0.87).

## DISCUSSION

In this one-sample Mendelian randomisation analysis, we found evidence of a causal link between vitamin D levels and diseases characterised by autoinflammation (i.e., by innate immune dysfunction at local sites), which was driven by the psoriasis as the most prevalent disease in this group. There was no statistical evidence of association between vitamin D and diseases characterised by autoimmunity (i.e., autoreactivity against native antigens predominantly due to adaptive immune dysfunction), although SLE may be one exception. We found no strong evidence for a plausible non-linear relationship between vitamin D and any outcome.

Vitamin D has been associated with numerous health outcomes in observational studies, which could not be replicated in randomised controlled trials [15]. Much of the trial evidence came from the landmark VITAL study, which found no difference between vitamin D supplement and placebo groups for a multitude of outcomes such as cancer and cardiovascular disease events [16], heart failure [17], atrial fibrillation [18], depression [19], body composition [20], falls [21], bone mineral density [22], fractures [23], frailty [24], knee pain [25], and biomarkers of inflammation [26]. However, the vitamin D group did have lower incidence of confirmed autoimmune diseases compared to placebo, but estimates included the null when including additional cases of probable autoimmune disease and/or excluding those with pre-randomisation autoimmune diseases [4]. Analyses of individual diseases (rheumatoid arthritis, polymyalgia rheumatica, psoriasis and a composite of the remainder) were all underpowered. Younger individuals and those with vitamin D deficiency (13%) were underrepresented. Taken together, VITAL provided long awaited randomised evidence, but much uncertainty remained in the context of multiple testing and regarding disease-specific or threshold effects.

Our results help to address these uncertainties. By leveraging much larger sample sizes of autoimmune diseases across the age spectrum, we show that vitamin D’s effect is likely to differ across individual diseases. We found evidence supporting a causal link between 25(OH)D and diseases characterised by autoinflammation but not autoimmunity. Genetic and clinical differences between these disease subgroups have been described previously [8], and is evident from the differential sex-predominance observed herein. The association with autoinflammatory diseases is almost certainly driven by psoriatic disease. The estimate from VITAL was directionally concordant but underpowered due to having only 38 psoriasis cases overall. We observed a similar association for psoriatic arthritis but with a wide confidence interval. The biological rationale for psoriasis is strong since topical vitamin D analogues are approved treatments. However, no high-quality trial evidence exists for oral vitamin D supplementation in reducing psoriasis risk. Among the diseases characterised by autoimmunity, SLE appears to be associated with vitamin D. This is consistent with prior observational evidence [27], but the 5 cases of SLE in VITAL could not power formal analyses. The associations between vitamin D and GCA and PMR should be interpreted with caution. Firstly, it is less biologically plausible that vitamin D should have opposing effects on two related diseases. Secondly, they were not replicated using the FinnGen data.

Taken together, our results suggest that vitamin D supplementation in those at risk of psoriasis or SLE may help reduce disease susceptibility, with no evidence indicating a threshold level. These results will also help inform future clinical trials that are needed to confirm our findings, for example, enriching populations with those at risk of psoriasis or SLE but not necessarily with vitamin D deficiency. The precise mechanism behind vitamin D’s effect on psoriasis and SLE cannot be derived from the current study. The VITAL trial found no benefit of vitamin D on IL6 or CRP [26] which are not the dominant inflammatory pathways in either disease. Future clinical trials should also focus on relevant mechanisms, e.g., involving T helper 17 and B cells.

The key strength of MR is that it is less susceptible to bias from confounding and reverse causation than conventional observational analyses. We selected instruments from genes relevant to vitamin D biology, which reduces potential bias from horizontal pleiotropy (i.e., arising from variants that do not have specific effects on vitamin D pathways). However, this important assumption cannot be empirically verified. Our use of biologically plausible variants may also explain why results differ from previous studies that adopted mechanism-agnostic variant-selection [28]. Misclassification of the outcome is possible when using diagnostic codes and self-report, which may increase risk of false negatives, particularly when sample sizes are small. Multiple testing is also of concern when considering individual diseases. However, we were able to replicate results for psoriasis and SLE using GWAS data derived from independent data with physician diagnosed cases. MR estimates the effect of subtle variations in lifelong exposure, thus results may not be comparable to therapeutic interventions. Our study only included participants of European ancestry. Future studies among other ethnic populations are needed to examine generalisability of the current findings, which is particularly relevant as skin colour influences vitamin D metabolism. Lastly, our data includes both prevalent and incident events; therefore, stratification into categories according to residual 25(OH)D concentration might be affected by reverse causation. However, genetic associations with disease outcomes within each of the strata will not be affected by reverse causation, as genotype is fixed from conception.

In conclusion, we found genetic evidence to support a causal link between 25(OH)D concentrations and autoinflammatory diseases such as psoriasis. These results have implications for the interpretation and design of vitamin D supplementation trials, and potential disease prevention strategies.

## Data Availability

UK Biobank data are available to all bona fide researchers for use in health-related research that is in the public interest. The application procedure is described at www.ukbiobank.ac.uk.

## Acknowledgements

We thank the participants of the UK Biobank study and the genome-wide association study consortia who made their summary statistics publicly available for this study. This work was supported by core funding from the: British Heart Foundation (RG/13/13/30194; RG/18/13/33946), BHF Cambridge CRE (RE/18/1/34212) and NIHR Cambridge Biomedical Research Centre (BRC-1215-20014). The views expressed are those of the author(s) and not necessarily those of the NIHR or the Department of Health and Social Care.

## Rights Retention Statement

For the purpose of open access, the author has applied a Creative Commons Attribution (CC BY) licence to any Author Accepted Manuscript version arising from this submission.

## Author contributions

All authors – SZ, AM, EG, HY, SB – have made substantial contributions to all of the following: (1) the conception and design of the study, or acquisition of data, or analysis and interpretation of data, (2) drafting the article or revising it critically for important intellectual content, (3) final approval of the version to be submitted. SZ takes responsibility for the integrity of the work as a whole, from inception to finished article.

## REFERENCES

1. Conrad N, Verbeke G, Molenberghs G, Goetschalckx L, Callender T, Cambridge G, et al. Autoimmune diseases and cardiovascular risk: a population-based study on 19 autoimmune diseases and 12 cardiovascular diseases in 22 million individuals in the UK. Lancet 2022;400:733–43.

2. Hayter SM, Cook MC. Updated assessment of the prevalence, spectrum and case definition of autoimmune disease. Autoimmun Rev 2012;11:754–65.

3. Kriegel MA, Manson JE, Costenbader KH. Does vitamin D affect risk of developing autoimmune disease?: a systematic review. Semin Arthritis Rheum 2011;40:512–531.e8.

4. Hahn J, Cook NR, Alexander EK, Friedman S, Walter J, Bubes V, et al. Vitamin D and marine omega 3 fatty acid supplementation and incident autoimmune disease: VITAL randomized controlled trial. BMJ 2022;376:e066452.

5. Bycroft C, Freeman C, Petkova D, Band G, Elliott LT, Sharp K, et al. The UK Biobank resource with deep phenotyping and genomic data. Nature 2018;562:203–9.

6. Astle WJ, Elding H, Jiang T, Allen D, Ruklisa D, Mann AL, et al. The Allelic Landscape of Human Blood Cell Trait Variation and Links to Common Complex Disease. Cell 2016;167:1415–1429.e19.

7. Mokry LE, Ross S, Ahmad OS, Forgetta V, Smith GD, Goltzman D, et al. Vitamin D and Risk of Multiple Sclerosis: A Mendelian Randomization Study. PLoS Med 2015;12:e1001866.

8. McGonagle D, McDermott MF. A Proposed Classification of the Immunological Diseases. PLOS Medicine 2006;3:e297.

9. McGonagle D, Aydin SZ, Gül A, Mahr A, Direskeneli H. ‘MHC-I-opathy’-unified concept for spondyloarthritis and Behçet disease. Nat Rev Rheumatol 2015;11:731–40.

10. Sofianopoulou E, Kaptoge SK, Afzal S, Jiang T, Gill D, Gundersen TE, et al. Estimating dose-response relationships for vitamin D with coronary heart disease, stroke, and all-cause mortality: observational and Mendelian randomisation analyses. The Lancet Diabetes & Endocrinology 2021;9:837–46.

11. Tian H, Mason AM, Liu C, Burgess S. Relaxing parametric assumptions for non-linear Mendelian randomization using a doubly-ranked stratification method [Internet]. 2022 [cited 2023 Feb 23];2022.06.28.497930. Available from: https://www.biorxiv.org/content/10.1101/2022.06.28.497930v1

12. Burgess S. Violation of the constant genetic effect assumption can result in biased estimates for non-linear Mendelian randomization [Internet]. 2022 [cited 2023 Feb 23];2022.10.26.22280570. Available from: https://www.medrxiv.org/content/10.1101/2022.10.26.22280570v1

13. Access results [Internet]. FinnGen [cited 2023 Feb 23];Available from: https://www.finngen.fi/en/access_results

14. Revez JA, Lin T, Qiao Z, Xue A, Holtz Y, Zhu Z, et al. Genome-wide association study identifies 143 loci associated with 25 hydroxyvitamin D concentration. Nat Commun 2020;11:1647.

15. Lucas A, Wolf M. Vitamin D and Health Outcomes: Then Came the Randomized Clinical Trials. JAMA 2019;322:1866–8.

16. Manson JE, Cook NR, Lee IM, Christen W, Bassuk SS, Mora S, et al. Vitamin D Supplements and Prevention of Cancer and Cardiovascular Disease. New England Journal of Medicine 2019;380:33–44.

17. Djoussé L, Cook NR, Kim E, Bodar V, Walter J, Bubes V, et al. Supplementation With Vitamin D and Omega-3 Fatty Acids and Incidence of Heart Failure Hospitalization. Circulation 2020;141:784–6.

18. Albert CM, Cook NR, Pester J, Moorthy MV, Ridge C, Danik JS, et al. Effect of Marine Omega-3 Fatty Acid and Vitamin D Supplementation on Incident Atrial Fibrillation: A Randomized Clinical Trial. JAMA 2021;325:1061–73.

19. Okereke OI, Reynolds CF III, Mischoulon D, Chang G, Vyas CM, Cook NR, et al. Effect of Long-term Vitamin D3 Supplementation vs Placebo on Risk of Depression or Clinically Relevant Depressive Symptoms and on Change in Mood Scores: A Randomized Clinical Trial. JAMA 2020;324:471–80.

20. Chou SH, Murata EM, Yu C, Danik J, Kotler G, Cook NR, et al. Effects of Vitamin D3 Supplementation on Body Composition in the VITamin D and OmegA-3 TriaL (VITAL). J Clin Endocrinol Metab 2021;106:1377–88.

21. LeBoff MS, Murata EM, Cook NR, Cawthon P, Chou SH, Kotler G, et al. VITamin D and OmegA-3 TriaL (VITAL): Effects of Vitamin D Supplements on Risk of Falls in the US Population. The Journal of Clinical Endocrinology & Metabolism 2020;105:2929–38.

22. LeBoff MS, Chou SH, Murata EM, Donlon CM, Cook NR, Mora S, et al. Effects of Supplemental Vitamin D on Bone Health Outcomes in Women and Men in the VITamin D and OmegA-3 TriaL (VITAL). J Bone Miner Res 2020;35:883–93.

23. LeBoff MS, Chou SH, Ratliff KA, Cook NR, Khurana B, Kim E, et al. Supplemental Vitamin D and Incident Fractures in Midlife and Older Adults. New England Journal of Medicine 2022;387:299–309.

24. Orkaby AR, Dushkes R, Ward R, Djousse L, Buring JE, Lee IM, et al. Effect of Vitamin D3 and Omega-3 Fatty Acid Supplementation on Risk of Frailty: An Ancillary Study of a Randomized Clinical Trial. JAMA Netw Open 2022;5:e2231206.

25. MacFarlane LA, Cook NR, Kim E, Lee IM, Iversen MD, Gordon D, et al. The Effects of Vitamin D and Marine Omega-3 Fatty Acid Supplementation on Chronic Knee Pain in Older US Adults: Results From a Randomized Trial. Arthritis Rheumatol 2020;72:1836–44.

26. Costenbader KH, MacFarlane LA, Lee IM, Buring JE, Mora S, Bubes V, et al. Effects of One Year of Vitamin D and Marine Omega-3 Fatty Acid Supplementation on Biomarkers of Systemic Inflammation in Older US Adults. Clin Chem 2019;65:1508–21.

27. Shoenfeld Y, Giacomelli R, Azrielant S, Berardicurti O, Reynolds JA, Bruce IN. Vitamin D and systemic lupus erythematosus - The hype and the hope. Autoimmunity Reviews 2018;17:19–23.

28. Bae SC, Lee YH. Vitamin D level and risk of systemic lupus erythematosus and rheumatoid arthritis: a Mendelian randomization. Clin Rheumatol 2018;37:2415–21.

