## Supplementary for "Associations between vitamin D and autoimmune diseases: Mendelian randomization analysis"

### Supplementary Table 1. Autoimmune disease and control outcome definitions

| **Outcome name** | **ICD-9 codes** | **ICD-10 codes** | **Self-report** | **N** |
| --- | --- | --- | --- | --- |
| Rheumatoid arthritis | 714.0, 714.1, 714.2, 714.81 | M05.0, M05.1, M05.2, M05.3, M05.8, M05.9, M06.0, M06.8, M06.9 | 1464 | 5172 |
| Systemic lupus erythematosus | 710.0 | M32.0, M32.1, M32.8, M32.9 | 1424 | 527 |
| Systemic sclerosis | 710.1 | M34.0, M34.1, M34.2, M34.8, M34.9 | 1384 | 201 |
| Sjogren’s syndrome | 710.2 | M35.0 | 1382 | 335 |
| Graves’ disease | 242.0 | E05.X | 1522 | 1798 |
| Hashimoto’s thyroiditis | 245.2 | E06.3 | - | 194 |
| Coeliac disease | 579.0 | K90.0 | 1456 | 2249 |
| Type 1 diabetes mellitus | 250.01 | E10 | 1222 | 341 |
| Primary biliary cholangitis | 571.6 | K74.3 | 1506 | 243 |
| Autoimmune hepatitis | 571.42 | K75.4 | - | 153 |
| Polymyalgia rheumatica | 725 | M35.3 | 1377 | 1780 |
| Giant cell arteritis | 446.5 | M31.5, M31.6 | 1376 | 436 |
| Polyarteritis nodosa | 446.0 | M30.0 | 1380 | 91 |
| Henoch-Schönlein Purpura | 287.0 | D69.0 | - | 47 |
| Granulomatosis with polyangiitis | 446.4 | M31.3 | 1378 | 62 |
| Eosinophilic granulomatosis with polyangiitis | - | M30.1 | - | 46 |
| Microscopic polyangiitis | - | M31.7 | 1379 | 19 |
| Mixed connective tissue disease | - | M35.1 | - | 41 |
| Antiphospholipid syndrome | - | D68.6 | 1564 | 28 |
| Dermato/polymyositis | 710.3, 710.4 | M33.1, M33.2, M33.9 | 1383 | 6 |
| Ankylosing spondylitis | 720.0 | M45 | 1313 | 1121 |
| Psoriatic arthritis | 696.0 | M07.0, M07.2, M07.3 | 1477 | 703 |
| Psoriasis | 696.1 | L40.0, L40.1, L40.2, L40.3, L40.4, L40.8, L40.9 | 1453 | 5120 |
| Crohn’s disease | 555.X | K50.X | 1462 | 1795 |
| Ulcerative colitis | 556.X | K51.X | 1463 | 3461 |
| Primary sclerosing cholangitis | 714.0, 714.1, 714.2, 714.81 | M05.0, M05.1, M05.2, M05.3, M05.8, M05.9, M06.0, M06.8, M06.9 | 1464 | 35 |
| Behcet's disease | 136.1 | M35.2 | - | 30 |
| Takayasu arteritis | 446.7 | M31.4 | - | 6 |
| Kawasaki disease | 446.1 | M30.3 | - | 1 |
| Multiple sclerosis | 340 | G35 | 1261 | 1504 |
| Osteoarthritis | 715.X | M15.0, M15.1, M15.2, M15.3, M15.4, M15.8, M15.9, M19.9 | 1465 | 31628 |

International Classification of Diseases (ICD) codes were used to match against conditions and cause of death, self-report codes were used to match against illness codes in UK Biobank data field 20002 (“non-cancer illness code, self-reported”). The .X notation means that any subcode is matched. Only diseases with n>100 were included in analyses.

### Supplementary Table 2. Mean values of 25(OH)D concentration (nmol/L) by month of blood draw

| **Month** | **Mean 25(OH)D concentration (nmol/L)** |
| --- | --- |
| January | 44.4 |
| February | 40.1 |
| March | 39.4 |
| April | 39.5 |
| May | 42.0 |
| June | 47.0 |
| July | 55.1 |
| August | 59.7 |
| September | 60.7 |
| October | 60.0 |
| November | 55.4 |
| December | 48.5 |

### Supplementary Table 3. List of genetic variants in the genetic risk score

| Chromosome: Position (hg19) | rsID | Effect allele | Other allele | Conditional association with 25(OH)D (nmol/L) |
| --- | --- | --- | --- | --- |
| 4:72617775 | rs1352846 | G | A | 0.172 |
| 4:72618334 | rs7041 | C | A | -0.045 |
| 4:72634343 | rs4694431 | T | C | -0.034 |
| 4:72770563 | rs139148694 | GTGCTTTTATCAA | G | 0.028 |
| 11:14339328 | rs16913816 | A | G | -0.031 |
| 11:14900931 | rs117913124 | A | G | 0.503 |
| 11:14912573 | rs117576073 | T | G | 0.246 |
| 11:14913575 | rs12794714 | A | G | 0.139 |
| 11:14913645 | rs202122669 | A | G | -0.615 |
| 11:14913900 | rs187639972 | C | G | -0.360 |
| 11:14941652 | rs117115472 | G | C | 0.148 |
| 11:71157867 | rs139168803 | A | G | -0.188 |
| 11:71158672 | rs12573951 | G | A | -0.045 |
| 11:71161063 | rs7928249 | G | A | -0.131 |
| 11:71180762 | rs549000212 | A | C | -0.364 |
| 11:71290740 | rs4081429 | C | A | 0.017 |
| 20:52714706 | rs6123359 | G | A | -0.026 |
| 20:52731402 | rs6127099 | T | A | 0.013 |
| 20:52735238 | rs35870583 | GT | G | 0.027 |
| 20:52737123 | rs2585442 | G | C | -0.025 |
| 20:52788925 | rs2762942 | A | G | -0.053 |

### Supplementary Table 4. Non-linear associations between 25(OHD) and each outcome across five quantiles.

| **Outcome quantile** | **Mean 25(OH)D** | **minimum 25(OH)D** | **Maximum 25(OH)D** | **OR** | **95% CI** | | **p value** |
| --- | --- | --- | --- | --- | --- | --- | --- |
| All autoimmune diseases 1 | 35.2 | 27.4 | 47.7 | 0.958 | 0.838 | 1.094 | 0.525 |
| All autoimmune diseases 2 | 45.2 | 33.2 | 58.6 | 0.856 | 0.746 | 0.983 | 0.027 |
| All autoimmune diseases 3 | 54.1 | 40.5 | 69.0 | 1.078 | 0.936 | 1.241 | 0.298 |
| All autoimmune diseases 4 | 63.9 | 48.2 | 80.4 | 0.903 | 0.787 | 1.036 | 0.147 |
| All autoimmune diseases 5 | 78.4 | 58.4 | 91.3 | 0.983 | 0.859 | 1.125 | 0.801 |
| Autoimmunity subgroup 1 | 35.2 | 27.4 | 47.7 | 0.975 | 0.816 | 1.164 | 0.777 |
| Autoimmunity subgroup 2 | 45.2 | 33.2 | 58.6 | 0.875 | 0.727 | 1.053 | 0.156 |
| Autoimmunity subgroup 3 | 54.1 | 40.5 | 69.0 | 1.025 | 0.848 | 1.238 | 0.798 |
| Autoimmunity subgroup 4 | 63.9 | 48.2 | 80.4 | 1.051 | 0.874 | 1.264 | 0.597 |
| Autoimmunity subgroup 5 | 78.4 | 58.4 | 91.3 | 1.012 | 0.850 | 1.205 | 0.891 |
| Autoinflammation subgroup 1 | 35.2 | 27.4 | 47.7 | 0.948 | 0.787 | 1.141 | 0.573 |
| Autoinflammation subgroup 2 | 45.2 | 33.2 | 58.6 | 0.822 | 0.680 | 0.995 | 0.044 |
| Autoinflammation subgroup 3 | 54.1 | 40.5 | 69.0 | 1.128 | 0.926 | 1.374 | 0.231 |
| Autoinflammation subgroup 4 | 63.9 | 48.2 | 80.4 | 0.765 | 0.631 | 0.927 | 0.006 |
| Autoinflammation subgroup 5 | 78.4 | 58.4 | 91.3 | 0.918 | 0.756 | 1.116 | 0.391 |
| Ankylosing spondylitis 1 | 35.2 | 27.4 | 47.7 | 1.213 | 0.647 | 2.271 | 0.547 |
| Ankylosing spondylitis 2 | 45.2 | 33.2 | 58.6 | 0.932 | 0.517 | 1.681 | 0.816 |
| Ankylosing spondylitis 3 | 54.1 | 40.5 | 69.0 | 0.906 | 0.498 | 1.650 | 0.747 |
| Ankylosing spondylitis 4 | 63.9 | 48.2 | 80.4 | 0.778 | 0.427 | 1.416 | 0.411 |
| Ankylosing spondylitis 5 | 78.4 | 58.4 | 91.3 | 0.997 | 0.560 | 1.775 | 0.991 |
| Giant cell arteritis 1 | 35.2 | 27.4 | 47.7 | 1.144 | 0.402 | 3.250 | 0.801 |
| Giant cell arteritis 2 | 45.2 | 33.2 | 58.6 | 0.368 | 0.138 | 0.979 | 0.045 |
| Giant cell arteritis 3 | 54.1 | 40.5 | 69.0 | 0.808 | 0.321 | 2.036 | 0.651 |
| Giant cell arteritis 4 | 63.9 | 48.2 | 80.4 | 0.499 | 0.209 | 1.189 | 0.117 |
| Giant cell arteritis 5 | 78.4 | 58.4 | 91.3 | 0.674 | 0.279 | 1.628 | 0.381 |
| Graves’ disease 1 | 35.2 | 27.4 | 47.7 | 1.030 | 0.660 | 1.608 | 0.896 |
| Graves’ disease 2 | 45.2 | 33.2 | 58.6 | 0.945 | 0.603 | 1.481 | 0.805 |
| Graves’ disease 3 | 54.1 | 40.5 | 69.0 | 0.829 | 0.518 | 1.329 | 0.437 |
| Graves’ disease 4 | 63.9 | 48.2 | 80.4 | 1.098 | 0.671 | 1.795 | 0.710 |
| Graves’ disease 5 | 78.4 | 58.4 | 91.3 | 0.992 | 0.592 | 1.662 | 0.976 |
| Psoriatic arthritis 1 | 35.2 | 27.4 | 47.7 | 0.806 | 0.415 | 1.565 | 0.525 |
| Psoriatic arthritis 2 | 45.2 | 33.2 | 58.6 | 0.542 | 0.262 | 1.123 | 0.099 |
| Psoriatic arthritis 3 | 54.1 | 40.5 | 69.0 | 2.200 | 0.967 | 5.006 | 0.060 |
| Psoriatic arthritis 4 | 63.9 | 48.2 | 80.4 | 0.731 | 0.336 | 1.594 | 0.431 |
| Psoriatic arthritis 5 | 78.4 | 58.4 | 91.3 | 0.750 | 0.341 | 1.650 | 0.474 |
| Polymyalgia rheumatica 1 | 35.2 | 27.4 | 47.7 | 0.932 | 0.554 | 1.568 | 0.792 |
| Polymyalgia rheumatica 2 | 45.2 | 33.2 | 58.6 | 1.803 | 1.059 | 3.068 | 0.030 |
| Polymyalgia rheumatica 3 | 54.1 | 40.5 | 69.0 | 1.175 | 0.722 | 1.912 | 0.518 |
| Polymyalgia rheumatica 4 | 63.9 | 48.2 | 80.4 | 1.263 | 0.799 | 1.995 | 0.317 |
| Polymyalgia rheumatica 5 | 78.4 | 58.4 | 91.3 | 1.164 | 0.752 | 1.801 | 0.496 |
| Rheumatoid arthritis 1 | 35.2 | 27.4 | 47.7 | 0.969 | 0.745 | 1.260 | 0.815 |
| Rheumatoid arthritis 2 | 45.2 | 33.2 | 58.6 | 0.740 | 0.559 | 0.979 | 0.035 |
| Rheumatoid arthritis 3 | 54.1 | 40.5 | 69.0 | 1.060 | 0.787 | 1.426 | 0.703 |
| Rheumatoid arthritis 4 | 63.9 | 48.2 | 80.4 | 1.058 | 0.793 | 1.410 | 0.702 |
| Rheumatoid arthritis 5 | 78.4 | 58.4 | 91.3 | 0.920 | 0.699 | 1.212 | 0.553 |
| Sjögren's syndrome 1 | 35.2 | 27.4 | 47.7 | 0.997 | 0.388 | 2.561 | 0.995 |
| Sjögren's syndrome 2 | 45.2 | 33.2 | 58.6 | 0.525 | 0.171 | 1.607 | 0.259 |
| Sjögren's syndrome 3 | 54.1 | 40.5 | 69.0 | 2.074 | 0.601 | 7.155 | 0.248 |
| Sjögren's syndrome 4 | 63.9 | 48.2 | 80.4 | 0.790 | 0.225 | 2.770 | 0.713 |
| Sjögren's syndrome 5 | 78.4 | 58.4 | 91.3 | 1.020 | 0.370 | 2.811 | 0.970 |
| Systemic lupus erythematosus 1 | 35.2 | 27.4 | 47.7 | 0.706 | 0.328 | 1.519 | 0.373 |
| Systemic lupus erythematosus 2 | 45.2 | 33.2 | 58.6 | 0.467 | 0.203 | 1.077 | 0.074 |
| Systemic lupus erythematosus 3 | 54.1 | 40.5 | 69.0 | 0.486 | 0.193 | 1.223 | 0.125 |
| Systemic lupus erythematosus 4 | 63.9 | 48.2 | 80.4 | 0.832 | 0.331 | 2.093 | 0.696 |
| Systemic lupus erythematosus 5 | 78.4 | 58.4 | 91.3 | 1.339 | 0.570 | 3.148 | 0.503 |
| Systemic sclerosis 1 | 35.2 | 27.4 | 47.7 | 1.763 | 0.500 | 6.222 | 0.378 |
| Systemic sclerosis 2 | 45.2 | 33.2 | 58.6 | 1.504 | 0.324 | 6.978 | 0.602 |
| Systemic sclerosis 3 | 54.1 | 40.5 | 69.0 | 2.028 | 0.529 | 7.779 | 0.303 |
| Systemic sclerosis 4 | 63.9 | 48.2 | 80.4 | 1.361 | 0.224 | 8.256 | 0.738 |
| Systemic sclerosis 5 | 78.4 | 58.4 | 91.3 | 0.362 | 0.091 | 1.438 | 0.149 |
| Autoimmune hepatitis 1 | 35.2 | 27.4 | 47.7 | 0.521 | 0.149 | 1.825 | 0.308 |
| Autoimmune hepatitis 2 | 45.2 | 33.2 | 58.6 | 3.382 | 0.491 | 23.277 | 0.216 |
| Autoimmune hepatitis 3 | 54.1 | 40.5 | 69.0 | 1.100 | 0.132 | 9.178 | 0.930 |
| Autoimmune hepatitis 4 | 63.9 | 48.2 | 80.4 | 0.186 | 0.041 | 0.847 | 0.030 |
| Autoimmune hepatitis 5 | 78.4 | 58.4 | 91.3 | 3.720 | 0.732 | 18.910 | 0.113 |
| Crohn’s disease 1 | 35.2 | 27.4 | 47.7 | 0.925 | 0.590 | 1.449 | 0.733 |
| Crohn’s disease 2 | 45.2 | 33.2 | 58.6 | 0.863 | 0.545 | 1.368 | 0.531 |
| Crohn’s disease 3 | 54.1 | 40.5 | 69.0 | 1.795 | 1.077 | 2.990 | 0.025 |
| Crohn’s disease 4 | 63.9 | 48.2 | 80.4 | 0.760 | 0.475 | 1.215 | 0.251 |
| Crohn’s disease 5 | 78.4 | 58.4 | 91.3 | 0.815 | 0.503 | 1.320 | 0.405 |
| Coeliac disease 1 | 35.2 | 27.4 | 47.7 | 1.249 | 0.790 | 1.976 | 0.341 |
| Coeliac disease 2 | 45.2 | 33.2 | 58.6 | 0.843 | 0.533 | 1.334 | 0.467 |
| Coeliac disease 3 | 54.1 | 40.5 | 69.0 | 1.243 | 0.787 | 1.961 | 0.351 |
| Coeliac disease 4 | 63.9 | 48.2 | 80.4 | 0.863 | 0.567 | 1.311 | 0.489 |
| Coeliac disease 5 | 78.4 | 58.4 | 91.3 | 1.108 | 0.773 | 1.588 | 0.575 |
| Hashimoto’s thyroiditis 1 | 35.2 | 27.4 | 47.7 | 1.060 | 0.267 | 4.213 | 0.934 |
| Hashimoto’s thyroiditis 2 | 45.2 | 33.2 | 58.6 | 2.121 | 0.489 | 9.199 | 0.315 |
| Hashimoto’s thyroiditis 3 | 54.1 | 40.5 | 69.0 | 1.083 | 0.281 | 4.172 | 0.908 |
| Hashimoto’s thyroiditis 4 | 63.9 | 48.2 | 80.4 | 1.164 | 0.224 | 6.064 | 0.857 |
| Hashimoto’s thyroiditis 5 | 78.4 | 58.4 | 91.3 | 0.271 | 0.070 | 1.053 | 0.059 |
| Primary biliary cholangitis 1 | 35.2 | 27.4 | 47.7 | 1.205 | 0.375 | 3.871 | 0.754 |
| Primary biliary cholangitis 2 | 45.2 | 33.2 | 58.6 | 0.391 | 0.091 | 1.678 | 0.206 |
| Primary biliary cholangitis 3 | 54.1 | 40.5 | 69.0 | 1.505 | 0.380 | 5.967 | 0.561 |
| Primary biliary cholangitis 4 | 63.9 | 48.2 | 80.4 | 1.930 | 0.484 | 7.696 | 0.352 |
| Primary biliary cholangitis 5 | 78.4 | 58.4 | 91.3 | 2.120 | 0.636 | 7.068 | 0.221 |
| Psoriasis 1 | 35.2 | 27.4 | 47.7 | 0.884 | 0.680 | 1.148 | 0.356 |
| Psoriasis 2 | 45.2 | 33.2 | 58.6 | 0.772 | 0.586 | 1.019 | 0.067 |
| Psoriasis 3 | 54.1 | 40.5 | 69.0 | 0.950 | 0.713 | 1.264 | 0.723 |
| Psoriasis 4 | 63.9 | 48.2 | 80.4 | 0.804 | 0.602 | 1.074 | 0.140 |
| Psoriasis 5 | 78.4 | 58.4 | 91.3 | 0.777 | 0.584 | 1.034 | 0.083 |
| Type 1 diabetes mellitus 1 | 35.2 | 27.4 | 47.7 | 0.672 | 0.252 | 1.794 | 0.428 |
| Type 1 diabetes mellitus 2 | 45.2 | 33.2 | 58.6 | 1.373 | 0.485 | 3.887 | 0.550 |
| Type 1 diabetes mellitus 3 | 54.1 | 40.5 | 69.0 | 0.846 | 0.283 | 2.529 | 0.765 |
| Type 1 diabetes mellitus 4 | 63.9 | 48.2 | 80.4 | 1.786 | 0.553 | 5.770 | 0.332 |
| Type 1 diabetes mellitus 5 | 78.4 | 58.4 | 91.3 | 1.745 | 0.506 | 6.021 | 0.378 |
| Ulcerative colitis 1 | 35.2 | 27.4 | 47.7 | 0.803 | 0.570 | 1.133 | 0.212 |
| Ulcerative colitis 2 | 45.2 | 33.2 | 58.6 | 0.902 | 0.640 | 1.272 | 0.557 |
| Ulcerative colitis 3 | 54.1 | 40.5 | 69.0 | 1.099 | 0.777 | 1.554 | 0.593 |
| Ulcerative colitis 4 | 63.9 | 48.2 | 80.4 | 0.739 | 0.532 | 1.025 | 0.070 |
| Ulcerative colitis 5 | 78.4 | 58.4 | 91.3 | 1.223 | 0.865 | 1.727 | 0.254 |

### Supplementary Table 5. Two-sample MR pleiotropy robust sensitivity analyses.

| **Outcome** | **Method** | **No. SNPs** | **OR** | **95% CI** | | **p value** |
| --- | --- | --- | --- | --- | --- | --- |
| Psoriasis | Inverse variance weighted | 2 | 0.518 | 0.278 | 0.963 | 0.038 |
| Systemic lupus erythematosus | Inverse variance weighted | 12 | 0.610 | 0.401 | 0.928 | 0.021 |
| Systemic lupus erythematosus | MR Egger | 12 | 0.506 | 0.220 | 1.163 | 0.140 |
| Systemic lupus erythematosus | Weighted median | 12 | 0.613 | 0.340 | 1.107 | 0.104 |
| Systemic lupus erythematosus | Weighted mode | 12 | 0.616 | 0.339 | 1.118 | 0.139 |
| Giant cell arteritis | Inverse variance weighted | 14 | 0.948 | 0.474 | 1.899 | 0.881 |
| Giant cell arteritis | MR Egger | 14 | 0.747 | 0.135 | 4.138 | 0.744 |
| Giant cell arteritis | Weighted median | 14 | 1.157 | 0.522 | 2.562 | 0.719 |
| Giant cell arteritis | Weighted mode | 14 | 1.213 | 0.539 | 2.732 | 0.648 |
| Polymyalgia rheumatica | Inverse variance weighted | 14 | 0.975 | 0.715 | 1.328 | 0.871 |
| Polymyalgia rheumatica | MR Egger | 14 | 0.755 | 0.362 | 1.575 | 0.468 |
| Polymyalgia rheumatica | Weighted median | 14 | 0.951 | 0.635 | 1.422 | 0.805 |
| Polymyalgia rheumatica | Weighted mode | 14 | 1.025 | 0.706 | 1.489 | 0.897 |
